# 30-Day Heart Failure Readmission and Post-Discharge Healthcare Use Among Older Australians: A Linked-Data Mixed-Methods Study

**DOI:** 10.64898/2026.10.09.26365178

**Authors:** Chang Sheng Leong, A Y M Alamgir Kabir, Margo Barr

## Abstract

**Background:** Heart failure (HF) is a major public health challenge in ageing populations and a leading cause of unplanned hospital readmissions. Biological, social, and economic factors contribute to recurrent hospitalisation, but gaps remain in understanding how these factors interact and patterns of healthcare use following hospitalisation.

**Methods:** We conducted an explanatory sequential mixed-methods study of patients aged ≥60 years with an HF hospital admission between 2016 and 2020 who completed the Sax Institute’s 45 and Up Study baseline questionnaire. Data were linked to Admitted Patient Data Collection, Medicare Benefits Schedule, and Pharmaceutical Benefits Scheme data. HF readmission within 30 days of the index admission was the outcome. Cox proportional hazards models estimated crude and adjusted hazard ratios (HRs and adjHRs) with 95% confidence intervals (CIs). Kaplan-Meier curves and log-rank tests compared survival between groups. Time-series plots mapped healthcare interactions over 1 year following the index admission.

**Results:** Among 4,252 patients with HF, 409 were readmitted within 30 days. Significant predictors included age ≥80 years (adjHR, 1.30; 95%CI, 1.01–1.67), 1 chronic condition (adjHR, 2.49; 95%CI, 1.41–4.40), ≥2 chronic conditions (adjHR, 1.75; 95%CI, 1.04–2.95), and alcohol consumption above national guidelines (adjHR, 1.45; 95%CI, 1.06–1.98). Time-series analysis indicated that participants with readmission had more frequent out-of-hospital health service contact following their initial HF admission than those without readmission.

**Conclusions:** Older age, chronic conditions, and high alcohol intake increased 30-day HF readmission risk. Differences in healthcare use after admission highlight opportunities for targeted and coordinated post-discharge care.

## Background

Heart failure (HF) is one of the most significant and rapidly growing challenges in global cardiovascular disease, driven by its substantial and increasing burden in the older adults. Approximately 64 million people globally are living with HF.^1,2^ Between 1990 and 2021, the prevalence of HF more than doubled, rising from 25.4 million to 55.5 million across all ages, while among people aged 65 and older, HF cases increased 2.5-fold, from 14.1 million to 36.2 million.^3^ Even though age-standardised incidence of HF has been declining in some high-income countries, the absolute burden of HF continues to grow because of population ageing, improved survival after myocardial infraction, and longer survival with HF itself once diagnosed.^4,5^ In Australia, the Australian Institute of Health and Welfare (AIHW) estimated 146,000 adults (0.7%) were living with HF in 2022–24, of whom almost two-thirds (91,600) were aged 65 years and over, with prevalence reaching 3.4% among those aged 75 years and older.^6^ As a result, HF has now become a leading and growing contributor to global morbidity, health care utilisation, and premature mortality, driven by its increasing prevalence and persistently poor prognosis.

As populations age, multimorbidity has become the norm rather than exception among older adults, and the coexistence of multiple chronic conditions is now major driver of health service use and cost.^7,8^ Within this broader landscape of chronic disease, HF is now one of the leading causes of hospitalisation in adults aged 65 years and older.^9^ In the United States, HF was the second leading cause of 30-day all cause hospital readmission among Medicare beneficiaries in 2020, behind only septicaemia.^10^ In Australia, cardiovascular disease is the leading broad cause of hospitalisation among adults aged 65 years and older, accounting for approximately one-fifth of all admissions in this age group, within which HF and cardiomyopathy is among the leading specific causes, ranking third behind coronary heart disease and atrial fibrillation. The rate of HF hospitalisations is nearly 20 times greater among those aged 75-79 years than those aged 45-49 years.^11^ The HF hospitalisation rate in Australia increased by a further 4.0% between 2014 and 2018.^12^ Additionally, AIHW estimated that the potentially preventable hospitalisations expenditure for HF in 2015-2016 was $597.3 million.^11^

For people with HF, readmissions contribute significantly to hospitalisations. For example, a study by Driscoll et al. (2020) estimated that approximately 24.0% of HF patients experience readmission within 30 days for any cause.^13^ Based on Su et al.’s (2019) review, the 30-day readmission rates range from 12.0% to 35.2% when examining readmissions with HF as a principal diagnosis.^14^ The readmission rate was 22 per 100 hospitalisations, excluding 1,103 index hospitalisations without 30-day follow-up.^15^ In 2017-18, there were 412,693 bed days attributed to HF, corresponding to 62,554 hospitalisations, representing 201 hospitalisations per 100,000 people of all ages (National rate).^11,16^ Chronic disease readmission rates in New South Wales (NSW) for 2015-2018 varied by condition and were highest for HF.^15^ Unplanned rehospitalisation within 30 days of discharge from an index HF admission was found to be associated with $604.4 million in direct hospital costs in Australia.^17^

Reducing avoidable readmissions improves health outcomes, patient safety, and the overall efficiency of the healthcare system. Australia’s aging population and rising rates of chronic disease are placing unprecedented pressure on individuals, communities, and the health system, with hospital readmission one of the many factors contributing to escalating healthcare costs. While risk factors for all-cause readmissions have been widely studied, research focused specifically on HF readmission remains limited and the few existing studies report inconsistent results.^18,19^ This study therefore aims to investigate the burden of HF-related hospital readmissions within 30 days of an index HF admission, and to identify factors associated with HF readmission among adults aged 60 years and older. It also seeks to investigate the healthcare interactions within one year following an index HF admission using a case-series approach.

## Methods

### Study Design and Population

We conducted an explanatory sequential mixed-methods design, comprising a quantitative retrospective cohort study and a qualitative time-series component.

Data for the quantitative study, were obtained from the Sax Institute’s 45 and Up Study, a large-scale cohort of 267,153 participants aged 45 years and above in NSW. Study participants were randomly sampled from the Services Australia Medicare enrolment database and invited to participate between 2005 and 2009 (representing roughly 11.0% of the NSW population aged 45 years and over). A baseline questionnaire was completed and provided written consent for follow-up data collection and linkage to electronic medical health records. The 45 and Up Study data were linked deterministically with the Medicare Benefits Schedule (MBS) and the Pharmaceutical Benefits Scheme (PBS) by the Sax Institute, while the NSW Admitted Patient Data Collection (APDC) and the NSW Registry of Births, Deaths and Marriages (RBDM) were linked probabilistically by the Centre for Health Record Linkage (CHeReL). Detailed study methods of the 45 and Up Study are available elsewhere.^20^ The study population comprised people aged 60 years and older who were identified as having HF hospital admission between 2016 and 2020. HF hospitalisation was defined as any hospital admission recorded in the APDC with a principal diagnosis of HF or hypertensive heart disease. HF was identified using International Statistical Classification of Diseases and Related Health Problems, 10th Revision, Australian Modification (ICD-10-AM) codes I50.1 and I50.9, while hypertensive heart disease was identified using codes I11, I13.0, and I13.2. Hypertensive heart disease was included in the heart failure definition because evidence indicates that some heart failure cases are coded under this category.^21^ The index admission for each patient was defined as their first HF hospitalisation during the study period (2016– 2020). Patient who died on the day of index hospital admission or within 30 days of follow up after the index hospitalisation were excluded from the cohort. In addition, patients identified as transfers of care from other facilities were excluded, defined as admissions in which HF was not recorded as the principal diagnosis at the time of initial admission.

### Outcome measures

The outcome of interest for the quantitative study was time to first HF readmission within 30 days following the index admission. HF readmission was defined as any hospitalisation to a NSW hospital within 30 days of discharge from the index admission with HF recorded as the principal diagnosis.

### Covariates

Socio-demographic and lifestyle variables were obtained from the baseline questionnaire of the 45 and Up Study and included age, sex, marital status, highest educational qualification, employment status, body mass index (BMI), smoking status, alcohol consumption, number of self-reported chronic conditions, self-reported quality of life, and social interaction. Social interaction was assessed using a social interaction index derived from four items of the Duke Social Support Index (DSSI): (1) spending time with friends or family members who do not live with the participant; (2) talking on the telephone with friends, relatives, or others; (3) attending meetings of social clubs, religious groups, or other organised groups; and (4) the number of people outside the participant’s household, but within one hour of travel, whom the participant felt they could depend on or felt very close to. Area-level socioeconomic status was determined using the Socio-Economic Indexes for Areas (SEIFA), and remoteness was classified using measures obtained from the Australian Bureau of Statistics (ABS).

### Quantitative Statistical analysis

HF readmission rate was calculated using person-time at risk as the denominator. Follow-up time was censored at the date of death or at completion of the 30-day follow-up period, whichever occurred first. Kaplan–Meier survival analysis was used to estimate and display the probability of remaining free from HF readmission during the 30-day period following the index hospitalisation. Kaplan–Meier survival curves were also used to compare the probability of remaining free from HF readmission across groups of key demographic and lifestyle characteristics, including age, sex, smoking status, and alcohol consumption. To identify the factors associated with HF readmission, univariate or multivariable Cox proportional regression models were used to estimate the crude and adjusted Hazard Ratio (HR and aHR respectively) and their 95% confidence interval (CI). Stata (StataCorp LLC. 2019. College Station, TX) statistical software was used to conduct all statistical analyses.

The research project received approval from two ethics committees: the NSW Population and Health Services Research Ethics Committee (Ref # 2019/ETH01682) for the data linkage and the University of NSW Human Research Ethics Committee, for the 45 and Up Study overall. The Sax Institute’s Secure Unified Research Environment (SURE) workspace provided a secure environment for conducting all analyses.

### Qualitative prospective time-series

For the prospective qualitative time-series analysis, a random subsample of 16 patients was selected, comprising 8 patients with HF–related admissions and 8 patients without

HF-related admissions. The index admission was defined using the same criteria as in the quantitative analysis. All selected patients were followed for 12 months (365 days) following the index admission.

During the follow-up period, longitudinal healthcare utilisation and vital events were identified using linked administrative datasets. Sequential events derived from hospital data included HF-related and non-HF-related readmissions, hospital discharges, and anaesthesia or procedure records. Events obtained from linked MBS data included general practitioner consultations, specialist consultations, and allied health services. Pharmacy visit was obtained from linked PBS data. Deaths were ascertained from the registries of deaths.

All sequential events were chronologically ordered and plotted over the 365-day follow-up period for both patient groups (HF-related admissions and non-HF-related admissions) to visualise individual care trajectories over time.

## Results

Among the participants from the 45 and Up Study who completed the baseline questionnaire, we ascertained 4,916 individuals aged 60 years and older with a principal diagnosis of HF or hypertensive heart disease who were admitted to hospital between 2016 and 2020. After implementing the exclusion criteria, the analytic cohort consisted of 4,252 HF patients (Figure 1).

**Figure 1:**
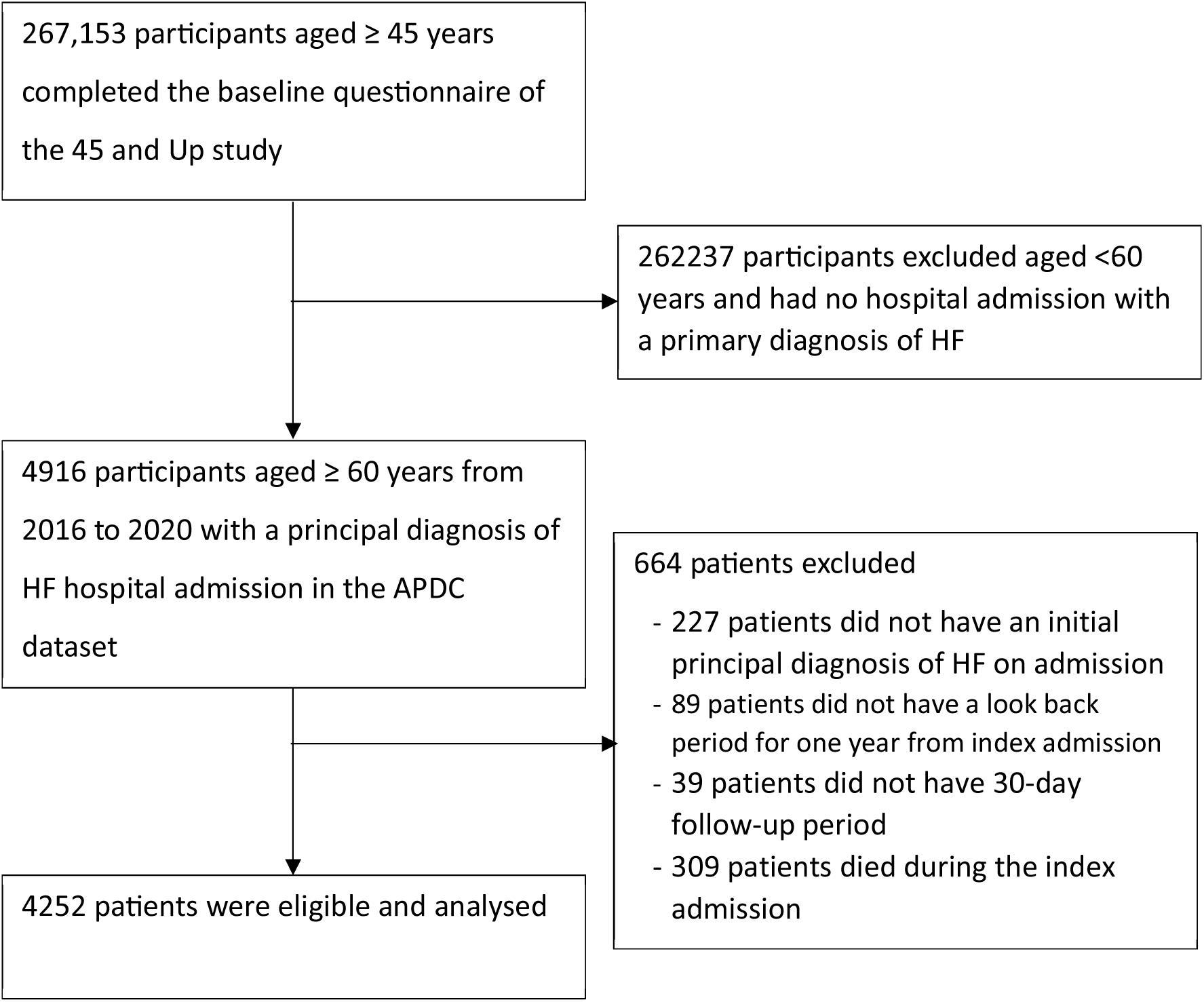
Assembly of analytic cohort selections.

Of these individuals, the predominant diagnosis category was congestive HF, accounting for 85% of the cohort, followed by left ventricular failure, which comprised 11% (Table 1). More than two-thirds of the study participants were aged 80 years and older, 56% were male, and about two-thirds were overweight or obese (Table 2). About half of the participants were current or ex-smoker (5% current and 44% ex-smoker), and 54% were alcohol drinkers with 16.8% drinking above the guideline. About 25% of the participants self-reported their quality of life was poor to fair and most (88%) were not doing any paid work. The multimorbidity (two or more chronic condition) prevalence was 78%.

**Table 1:**
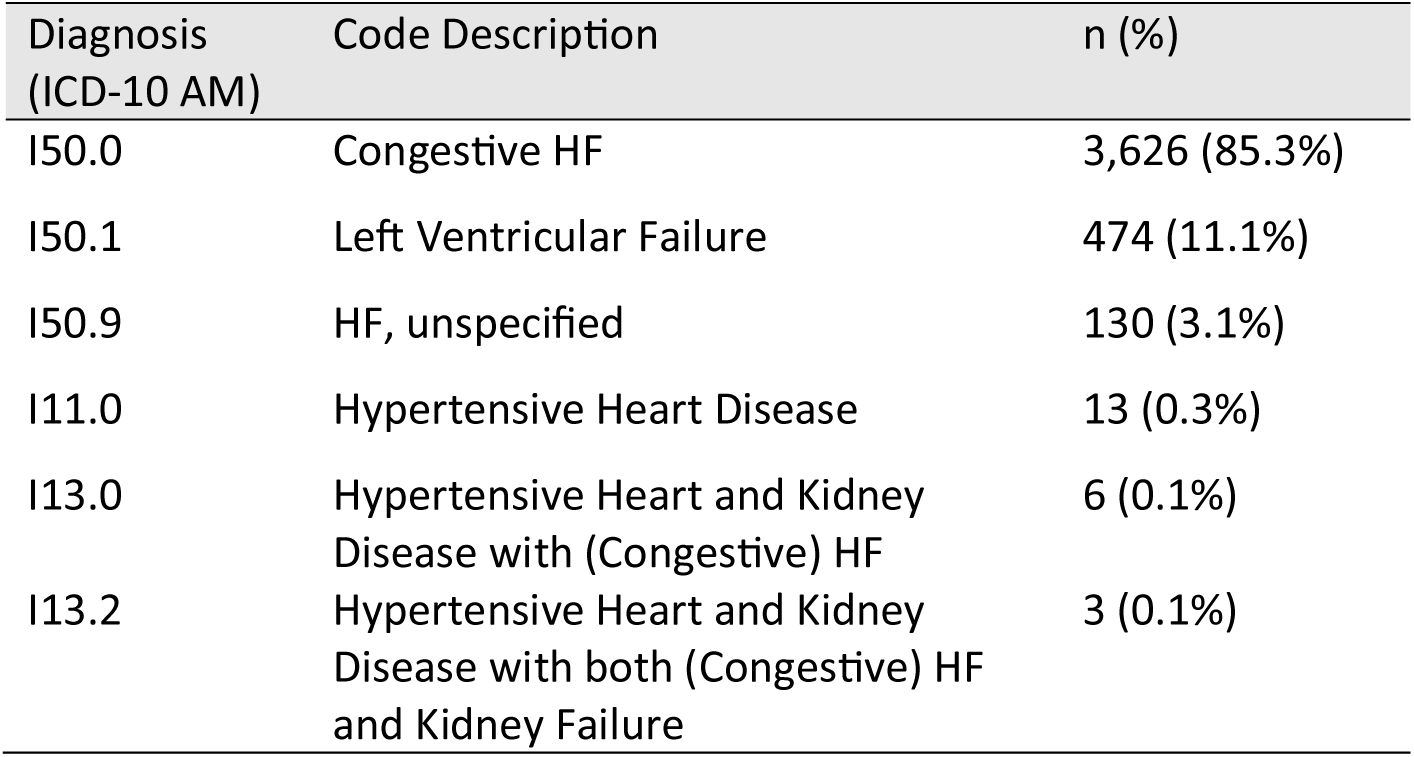
Analytic Cohort Composition According to ICD-10-AM Diagnosis Codes.

| Diagnosis (ICD-10 AM) | Code Description | n (%) |
| --- | --- | --- |
| I50.0 | Congestive HF | 3,626 (85.3%) |
| I50.1 | Left Ventricular Failure | 474 (11.1%) |
| I50.9 | HF, unspecified | 130 (3.1%) |
| I11.0 | Hypertensive Heart Disease | 13 (0.3%) |
| I13.0 | Hypertensive Heart and Kidney Disease with (Congestive) HF | 6 (0.1%) |
| I13.2 | Hypertensive Heart and Kidney Disease with both (Congestive) HF and Kidney Failure | 3 (0.1%) |

**Table 2:** Characteristics of the study participants.

| **Variable** | **No of participants** | **%** |
| --- | --- | --- |
| **Age** |  |  |
| 60 -79 years | 1,401 | 32.9 |
| 80 years and over | 2,851 | 67.1 |
| **Sex** |  |  |
| Male | 2,381 | 56.0 |
| Female | 1,871 | 44.0 |
| **BMI** |  |  |
| Underweight | 512 | 12.0 |
| Normal Weight | 1,048 | 24.6 |
| Overweight and Obese | 2,692 | 63.3 |
| **Number of Self-Reported Chronic Conditions** |  |  |
| 0 | 383.0 | 9.0 |
| 1 | 561 | 13.2 |
| ≥2 | 3,308 | 77.8 |
| **Smoking Status** |  |  |
| Never smoker | 2,186 | 51.4 |
| Ex-smoker | 1,866 | 43.9 |
| Current smoker | 200 | 4.7 |
| **Alcohol Intake** |  |  |
| None | 1,923 | 46.1 |
| Within guideline | 1,547 | 37.1 |
| Above guideline | 702 | 16.8 |
| **Self-Rated Quality of Life** |  |  |
| Poor - Fair | 1,033 | 25.3 |
| Good and above | 3,044 | 74.7 |
| **Duke Social Support Index (Social Interaction Score)** |  |  |
| DSSI 4 to 6 | 487 | 12.1 |
| DSSI 7 to 9 | 2,067 | 51.4 |
| DSSI 10 to 12 | 1,466 | 36.5 |
| **Marital Status** |  |  |
| Single/ Widowed/ Divorced/Separated | 1,697 | 40.1 |
| Married/De facto | 2,540 | 59.9 |
| **Highest Educational Qualification** |  |  |
| Up to high school | 2,359 | 56.9 |
| Up to Certificate or Diploma | 1,299 | 31.3 |
| University Degree or higher | 491 | 11.8 |
| **Employment status** |  |  |
| Employed | 509 | 12.0 |
| Not employed | 3,743 | 88.0 |
| **Healthcare Coverage** |  |  |
| None | 520 | 12.4 |
| Private Health Insurance/ Department of Veteran Affairs Card/Health Care Card | 3,663 | 87.6 |
| **Remoteness** |  |  |
| Urban | 2,316 | 55.1 |
| Regional and beyond | 1,888 | 44.9 |
| **SIEFA** |  |  |
| Most Disadvantage (Deciles 1 to 3) | 1,653 | 39.6 |
| Middle (Deciles 4 to 7) | 1,526 | 36.6 |
| Most Advantage (Deciles 8 to 10) | 990 | 23.7 |

### Readmission

Among 4,252 HF patients, 409 experienced a first HF readmission within 30 days of the index admission, corresponding to a rate of 1.3 readmissions per person-year, which was statistically significant (Rate: 1.3; 95% CI: 1.16–1.40; Table 3 and Figure 2).

**Table 3:** HF readmission rate within 30 days of index HF admission.

| Measure | Value |
| --- | --- |
| No of patients (a) | 4252 |
| Total person-days (b) | 117,067 |
| Total person-years (c) | 320.51 |
| No of HF-readmission within 30 days from index HF admission (d) | 409 |
| Rate of readmission per person-year (95% CI) (d/c) | 1.3 (1.16, 1.40) |

**Figure 2:**
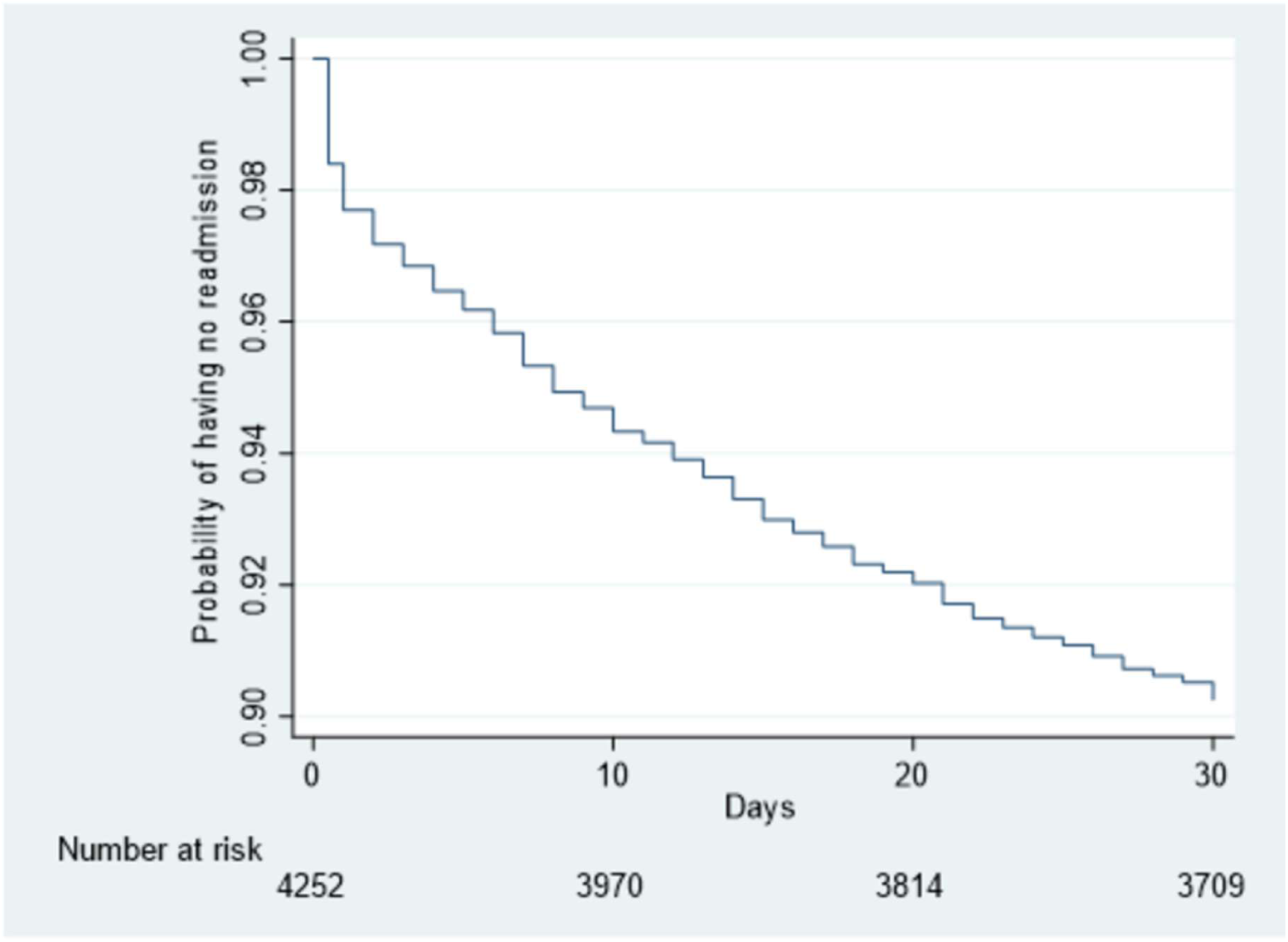
Kaplan-Meier survival curve for HF.

The readmission rate was higher among older patients (aged ≥80 years) compared to those aged 60–79 years (Table 4 and Figure 3). Male patients exhibited a higher readmission rate than their female counterparts. When stratified by the number of reported chronic conditions, patients with one chronic condition had the highest readmission rate, followed by those with two or more conditions, whereas patients with no chronic conditions had the lowest rate.

**Figure 3:**
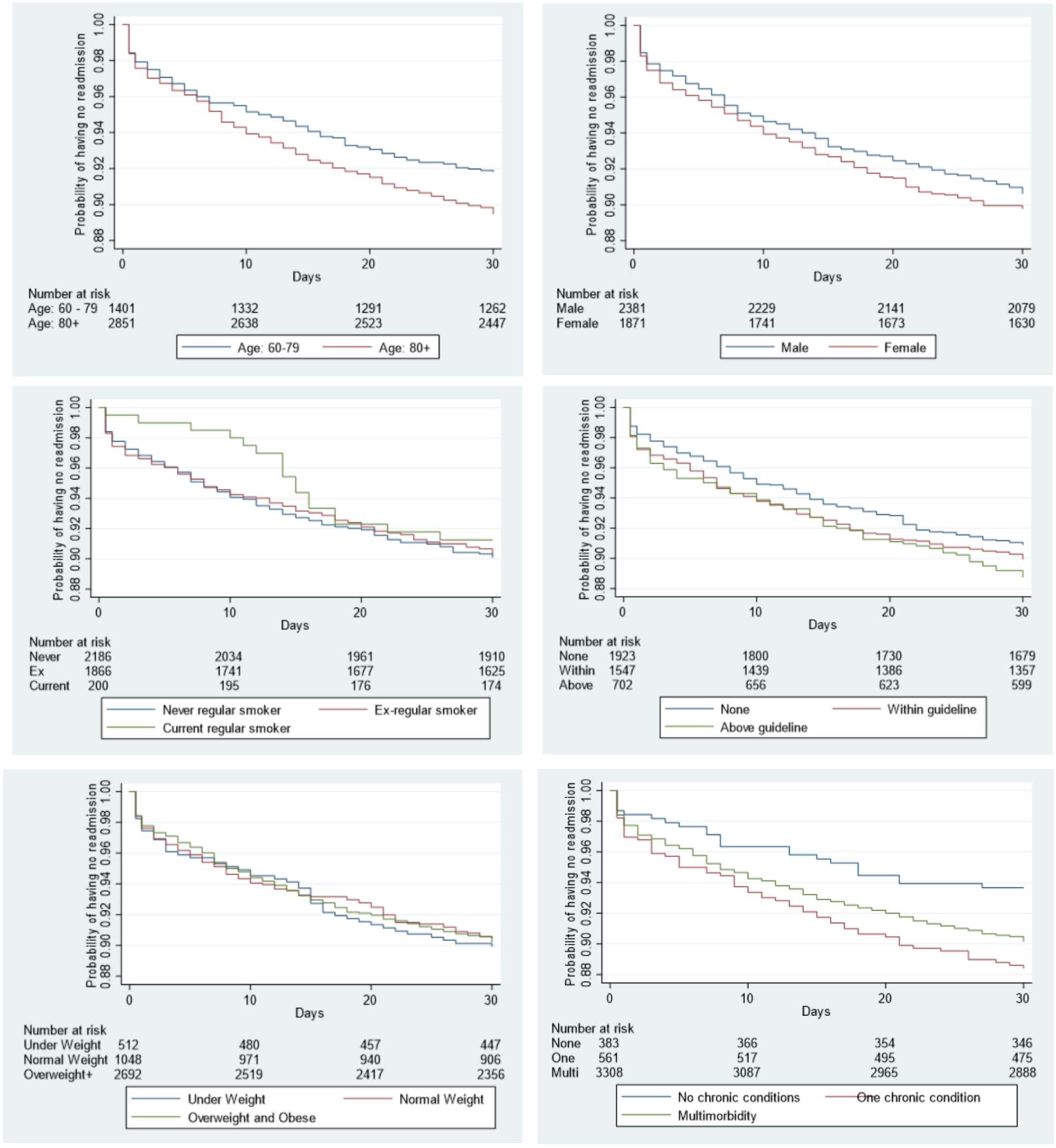
Kaplan-Meier curve for HF readmission over a 30-day period following discharge from an index HF admission by age, sex, smoking status, alcohol consumption, obesity and number of self-reported chronic conditions.

**Table 4:**
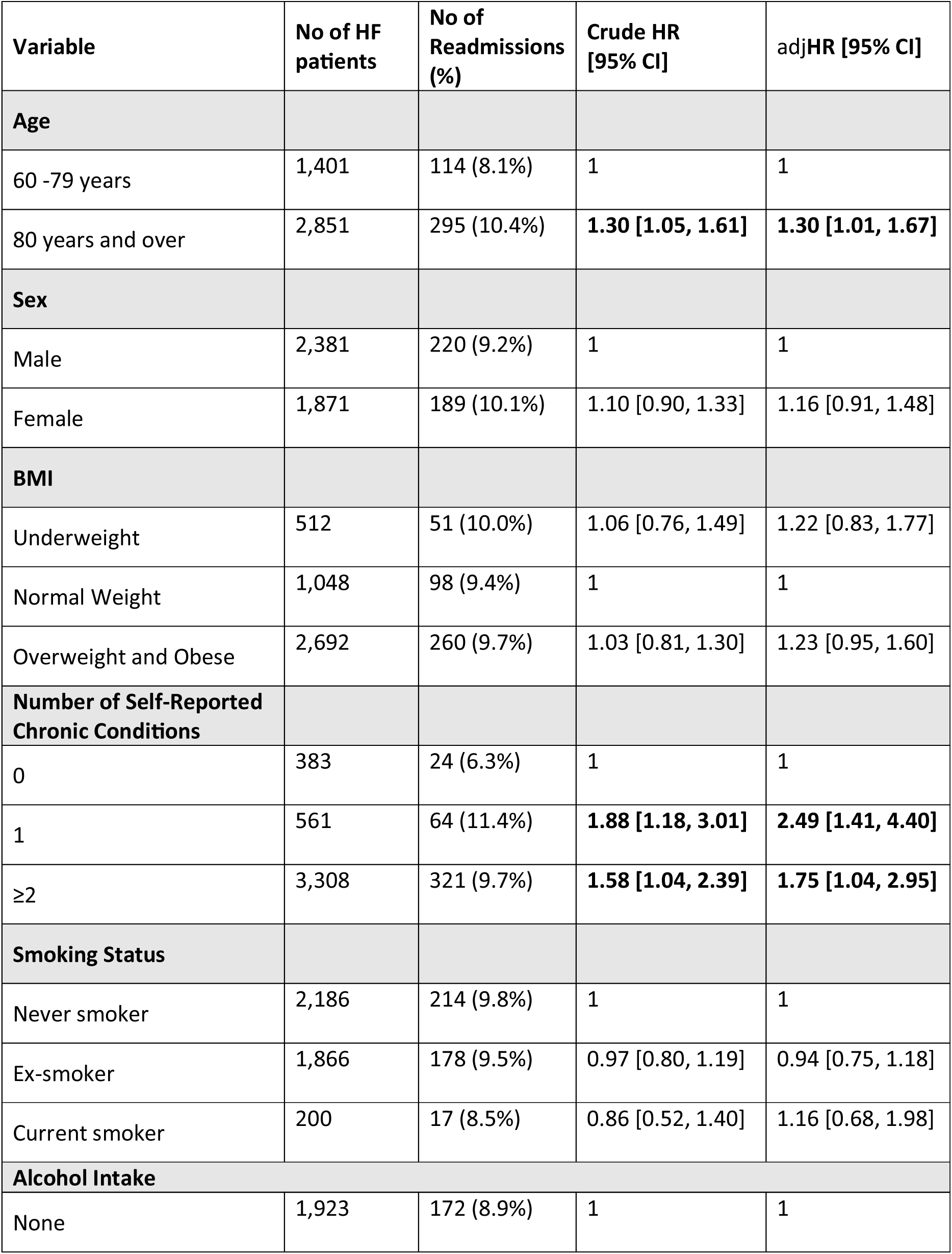
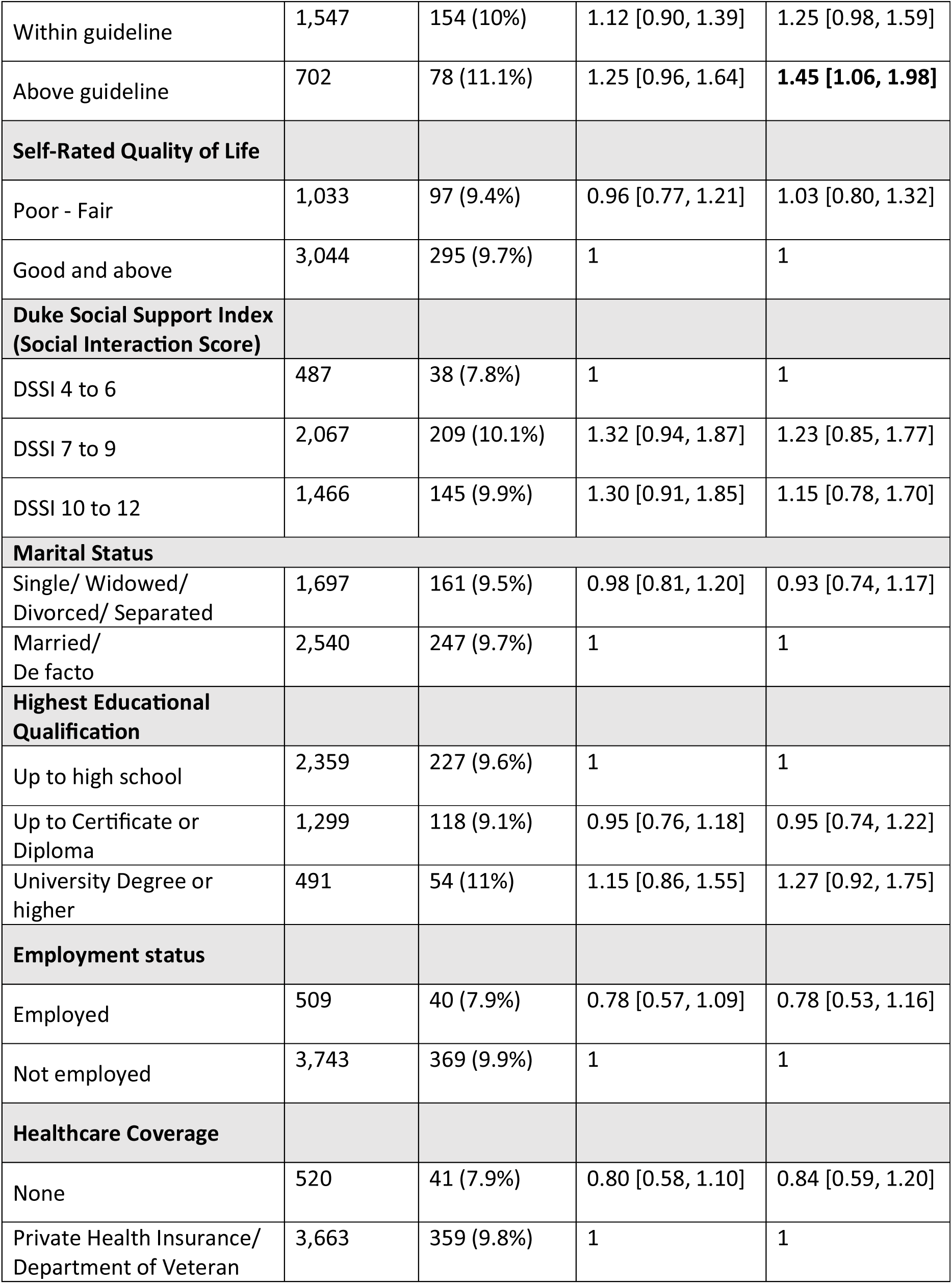
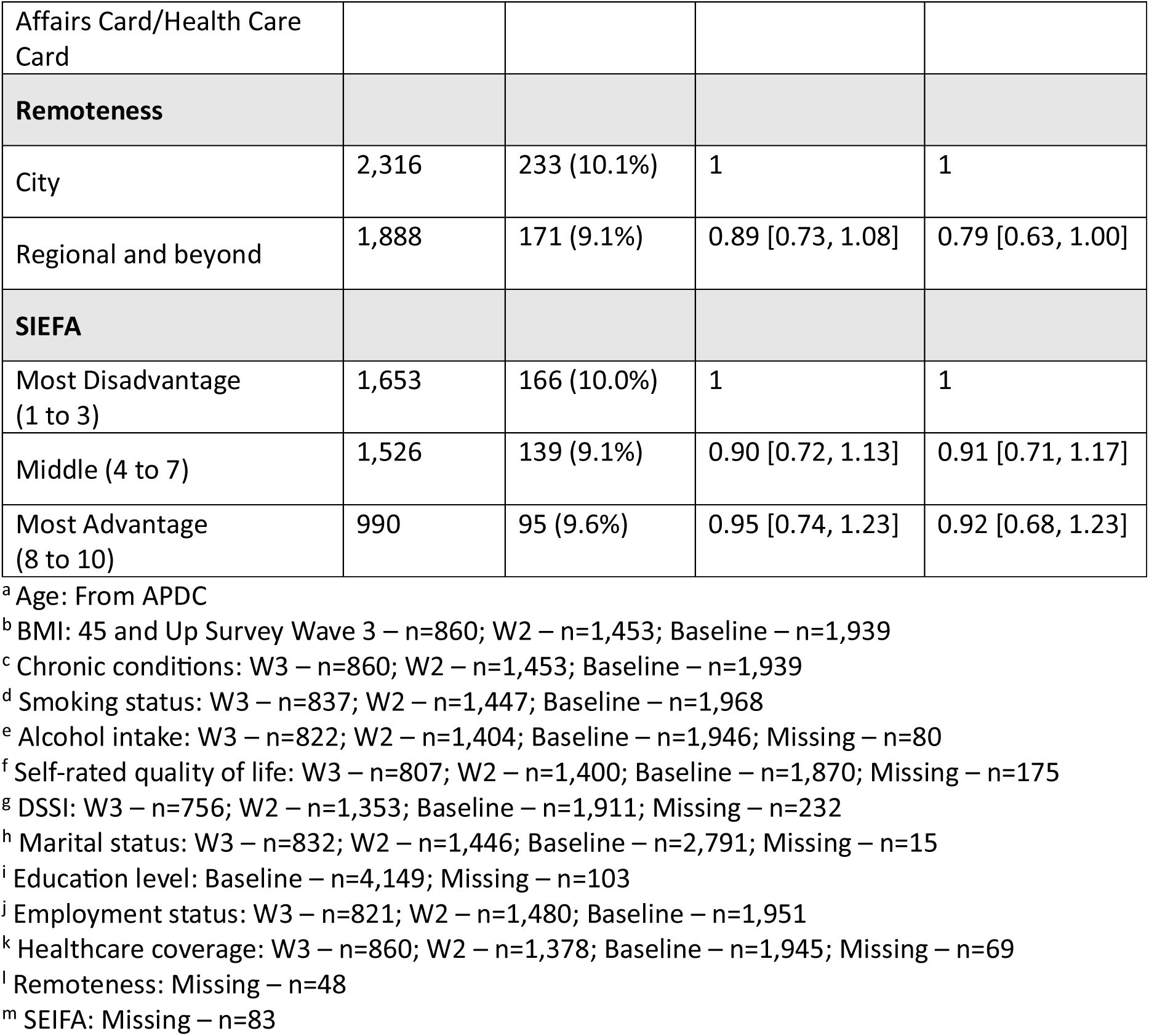
Factors associated with heart failure readmission within 30 days from the index heart failure hospital admission.

| Variable | No of HF patients | No of Readmissions (%) | Crude HR [95% CI] | adjHR [95% CI] |
| --- | --- | --- | --- | --- |
| **Age** |  |  |  |  |
| 60 -79 years | 1,401 | 114 (8.1%) | 1 | 1 |
| 80 years and over | 2,851 | 295 (10.4%) | **1.30 [1.05, 1.61]** | **1.30 [1.01, 1.67]** |
| **Sex** |  |  |  |  |
| Male | 2,381 | 220 (9.2%) | 1 | 1 |
| Female | 1,871 | 189 (10.1%) | 1.10 [0.90, 1.33] | 1.16 [0.91, 1.48] |
| **BMI** |  |  |  |  |
| Underweight | 512 | 51 (10.0%) | 1.06 [0.76, 1.49] | 1.22 [0.83, 1.77] |
| Normal Weight | 1,048 | 98 (9.4%) | 1 | 1 |
| Overweight and Obese | 2,692 | 260 (9.7%) | 1.03 [0.81, 1.30] | 1.23 [0.95, 1.60] |
| **Number of Self-Reported Chronic Conditions** |  |  |  |  |
| 0 | 383 | 24 (6.3%) | 1 | 1 |
| 1 | 561 | 64 (11.4%) | **1.88 [1.18, 3.01]** | **2.49 [1.41, 4.40]** |
| ≥2 | 3,308 | 321 (9.7%) | **1.58 [1.04, 2.39]** | **1.75 [1.04, 2.95]** |
| **Smoking Status** |  |  |  |  |
| Never smoker | 2,186 | 214 (9.8%) | 1 | 1 |
| Ex-smoker | 1,866 | 178 (9.5%) | 0.97 [0.80, 1.19] | 0.94 [0.75, 1.18] |
| Current smoker | 200 | 17 (8.5%) | 0.86 [0.52, 1.40] | 1.16 [0.68, 1.98] |
| **Alcohol Intake** |  |  |  |  |
| None | 1,923 | 172 (8.9%) | 1 | 1 |

| **Variable** | **No of HF patients** | **No of Readmissions (%)** | **Crude HR [95% CI]** | **adjHR [95% CI]** |
| --- | --- | --- | --- | --- |
| Within guideline | 1,547 | 154 (10%) | 1.12 [0.90, 1.39] | 1.25 [0.98, 1.59] |
| Above guideline | 702 | 78 (11.1%) | 1.25 [0.96, 1.64] | **1.45 [1.06, 1.98]** |
| **Self-Rated Quality of Life** |  |  |  |  |
| Poor - Fair | 1,033 | 97 (9.4%) | 0.96 [0.77, 1.21] | 1.03 [0.80, 1.32] |
| Good and above | 3,044 | 295 (9.7%) | 1 | 1 |
| **Duke Social Support Index (Social Interaction Score)** |  |  |  |  |
| DSSI 4 to 6 | 487 | 38 (7.8%) | 1 | 1 |
| DSSI 7 to 9 | 2,067 | 209 (10.1%) | 1.32 [0.94, 1.87] | 1.23 [0.85, 1.77] |
| DSSI 10 to 12 | 1,466 | 145 (9.9%) | 1.30 [0.91, 1.85] | 1.15 [0.78, 1.70] |
| **Marital Status** |  |  |  |  |
| Single/ Widowed/ Divorced/ Separated | 1,697 | 161 (9.5%) | 0.98 [0.81, 1.20] | 0.93 [0.74, 1.17] |
| Married/ De facto | 2,540 | 247 (9.7%) | 1 | 1 |
| **Highest Educational Qualification** |  |  |  |  |
| Up to high school | 2,359 | 227 (9.6%) | 1 | 1 |
| Up to Certificate or Diploma | 1,299 | 118 (9.1%) | 0.95 [0.76, 1.18] | 0.95 [0.74, 1.22] |
| University Degree or higher | 491 | 54 (11%) | 1.15 [0.86, 1.55] | 1.27 [0.92, 1.75] |
| **Employment status** |  |  |  |  |
| Employed | 509 | 40 (7.9%) | 0.78 [0.57, 1.09] | 0.78 [0.53, 1.16] |
| Not employed | 3,743 | 369 (9.9%) | 1 | 1 |
| **Healthcare Coverage** |  |  |  |  |
| None | 520 | 41 (7.9%) | 0.80 [0.58, 1.10] | 0.84 [0.59, 1.20] |
| Private Health Insurance/ Department of Veteran | 3,663 | 359 (9.8%) | 1 | 1 |
| Affairs Card/Health Care Card |  |  |  |  |
| **Remoteness** |  |  |  |  |
| City | 2,316 | 233 (10.1%) | 1 | 1 |
| Regional and beyond | 1,888 | 171 (9.1%) | 0.89 [0.73, 1.08] | 0.79 [0.63, 1.00] |
| **SIEFA** |  |  |  |  |
| Most Disadvantage (1 to 3) | 1,653 | 166 (10.0%) | 1 | 1 |
| Middle (4 to 7) | 1,526 | 139 (9.1%) | 0.90 [0.72, 1.13] | 0.91 [0.71, 1.17] |
| Most Advantage (8 to 10) | 990 | 95 (9.6%) | 0.95 [0.74, 1.23] | 0.92 [0.68, 1.23] |

<sup>a</sup> Age: From APDC
<sup>b</sup> BMI: 45 and Up Survey Wave 3 – n=860; W2 – n=1,453; Baseline – n=1,939
<sup>c</sup> Chronic conditions: W3 – n=860; W2 – n=1,453; Baseline – n=1,939
<sup>d</sup> Smoking status: W3 – n=837; W2 – n=1,447; Baseline – n=1,968
<sup>e</sup> Alcohol intake: W3 – n=822; W2 – n=1,404; Baseline – n=1,946; Missing – n=80
<sup>f</sup> Self-rated quality of life: W3 – n=807; W2 – n=1,400; Baseline – n=1,870; Missing – n=175
<sup>g</sup> DSSI: W3 – n=756; W2 – n=1,353; Baseline – n=1,911; Missing – n=232
<sup>h</sup> Marital status: W3 – n=832; W2 – n=1,446; Baseline – n=2,791; Missing – n=15
<sup>i</sup> Education level: Baseline – n=4,149; Missing – n=103
<sup>j</sup> Employment status: W3 – n=821; W2 – n=1,480; Baseline – n=1,951
<sup>k</sup> Healthcare coverage: W3 – n=860; W2 – n=1,378; Baseline – n=1,945; Missing – n=69
<sup>l</sup> Remoteness: Missing – n=48
<sup>m</sup> SEIFA: Missing – n=83

### Risk factors of HF readmission

Factors associated with 30-day readmission among HF patients are presented in Table 4. Patients aged ≥80 years associated with a 30% higher risk of readmission compared to those aged 60–79 years (adjHR: 1.30; 95% CI: 1.01–1.67). The number of self-reported chronic conditions was significantly associated with HF readmission. Compared with patients with no chronic conditions, those with one chronic condition associated with a 2.49-fold higher risk (adjHR: 2.49; 95% CI: 1.41–4.40), while those with two or more chronic conditions had a 1.75-fold increased risk (adjHR: 1.75; 95% CI: 1.04–2.95). Patients who consumed alcohol above recommended guidelines associated with a 45% higher risk of HF readmission (adjHR: 1.45; 95% CI: 1.06–1.98). In contrast, patients residing in regional areas were associated with a 21% lower risk of readmission compared to those living in metropolitan areas (adjHR: 0.79; 95% CI: 0.63–1.00). There was no statistical difference for the other factors considered.

### Qualitative Time-Series

#### Participant selection and characteristics

Age, chronic conditions and alcohol intake were identified as key covariates and were explored further through a participant-level descriptive analysis using the available linked data. Sixteen participants were randomly selected to illustrate these characteristics and their subsequent health service use and outcomes: eight participants without HF readmissions (IDs 1–8) and eight participants with HF readmissions (IDs 9–16). Among participants without HF readmissions (IDs 1–8), four (IDs 1, 2, 5 and 6) were aged 60–79 years and four (IDs 3, 4, 7 and 8) were aged 80 years or older. Participants 2, 4, 6 and 8 had one or more self-reported chronic conditions and reported alcohol consumption above the recommended guidelines. Participants 1, 3, 5 and 7 had no self-reported chronic conditions and either did not consume alcohol or reported consumption within the recommended guidelines. There were equal numbers of males and females. Among participants with HF readmissions (IDs 9–16), four (IDs 9, 10, 13 and 14) were aged 60–79 years and four (IDs 11, 12, 15 and 16) were aged 80 years or older. Participants 10, 12, 14 and 16 had one or more self-reported chronic conditions and reported alcohol consumption above the recommended guidelines. Participants 9, 11, 13 and 15 had no self-reported chronic conditions and either did not consume alcohol or reported consumption within the recommended guidelines. Again, there were equal numbers of males and females.

#### Hospital readmissions and mortality

The time series for all participants began with an index hospital admission for which HF was the principal diagnosis. Figures 4 presents participant-level timelines of hospital admissions, hospital discharge and deaths over the 365 days following the index admission. Different symbols represent different types of clinical events (as squares), as defined in the figure legend. As shown in Figure 4, among participants without HF readmissions (IDs 1–8), participants 1–5 and 8 subsequently had readmissions for other causes, while participants 6 and 7 had no further hospital admissions. Participants 2 and 4 died within one year of the index admission. Both had one or more self-reported chronic conditions and reported alcohol consumption above the recommended guidelines. All participants with HF readmissions (IDs 9–16) had at least one subsequent HF readmission. Six participants (IDs 9–12, 15 and 16) also had readmissions for other causes. Three participants died within one year of the index admission; two had one or more self-reported chronic conditions and reported alcohol consumption above the recommended guidelines. Some older participants had multiple HF readmissions, particularly participant 15.

**Figure 4:**
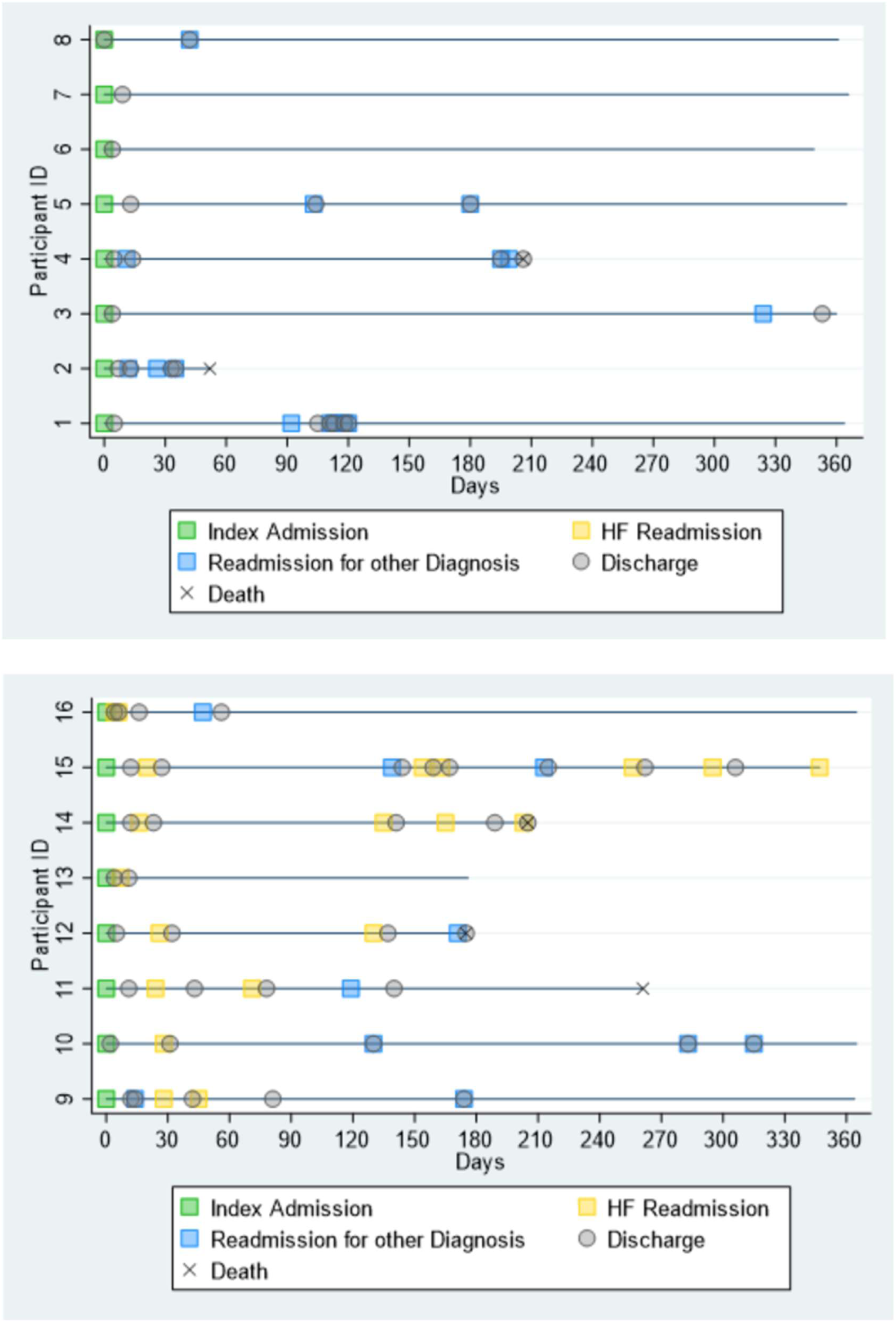
One-year time series of participants without (1-8) and with (9-16) HF readmissions using APDC administrative data.

#### Out-of-hospital health service use

Figure 5 presents participant-level timelines of hospital admissions, deaths as well as interactions with out-of-hospital health services over the 365 days following the index admission. Different symbols represent different types of clinical events (as squares) and out-of-hospital health service contact (as triangles) and prescriptions (as diamonds) as defined in the figure legend. No clear patterns were observed between out-of-hospital health service use and the characteristics examined, including age, number of chronic conditions or alcohol intake, or between out-of-hospital contacts and the number of hospital readmissions. However, participants with HF readmissions (IDs 9–16) appeared to have more frequent contact with out-of-hospital health services than participants without HF readmissions (IDs 1–8).

**Figure 5:**
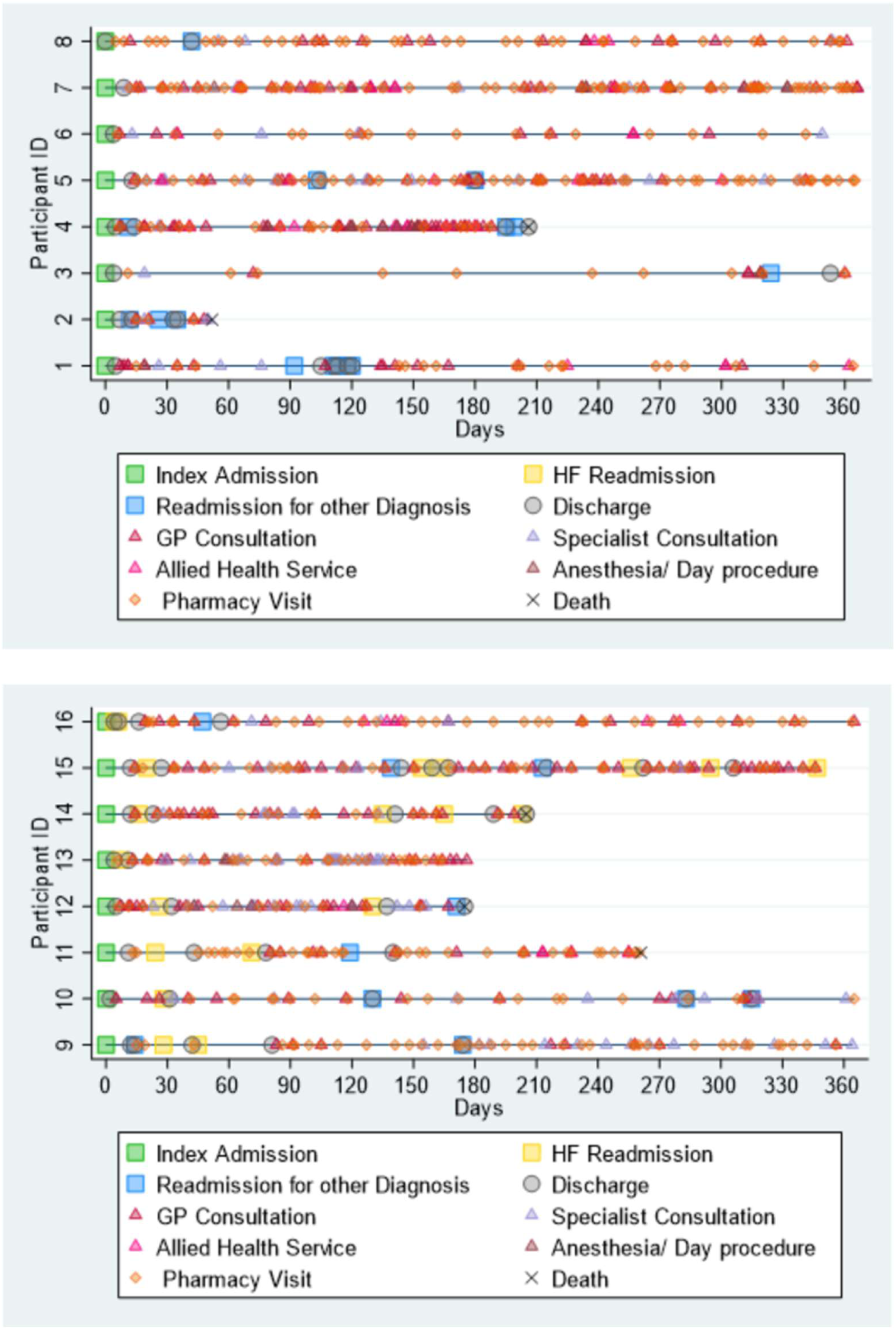
One-year time series of participants without (1-8) and with (9-16) HF readmissions using linked population-based data: APDC, MBS, and PBS.

## Discussion

This was a large population-based data linkage retrospective cohort study examining the incidence of HF readmission within 30 days of index HF hospital admission and its risk factors. We found an incidence rate 1.3 HF readmissions per 100,000 person-years within 30 days of index HF admission. Age ≥80 years was fond to be associated with a 30% increased risk of 30-day HF readmission, and having a self-reported chronic condition was associated with a 2.49-fold increased risk. Alcohol consumption above the recommended guidelines was associated with a 45% increased risk, while living in a regional area was associated with a 21% lower risk of 30-day HF readmission.

It is difficult to compare the findings of our study with those of other studies because of variations in the data sources, standardisation methods (30 days versus 90 days versus 1-year readmission), and definitions of HF and readmission (all-cause versus HF-only) used. Nevertheless, in a one-year HF readmission study, comorbidities were also found to be significantly associated with HF readmission risk (HR=1.53, 95% CI: 1.10 to 2.11, p=0.011).^22^ Similar to our paper, a 30-day HF readmission study by Schjødt et al. (2019) on education levels revealed no significant risk in low-level education (adjHR=1.11, 95% CI: 0.86 to 1.42) and Mid-level education (adjHR=1.06, 95% CI: 0.83 to 1.36).^23^ This was the same for education and all-cause readmissions in Sterling et al.’s REGARDS study (HR=1.08, 95%CI: 0.75-1.56). Likewise, the REGARDS study also found no statistical significance for HF readmission risk when examining rural residence (HR: 1.20, CI:0.72–1.99) and social isolation (HR 1.30, CI:0.84–2.00).^24^

Our study did not find any significant association between social support (DSSI) or marital status and HF readmission. However, prior research by Rodríguez-Artalejo et al. in 2006 did discover a link between social support and HF readmission, with patients having moderate (adjHR=1.87; 95% CI: 1.06-3.29; p < .05) or low levels (adjHR 1.98; 95% CI: 1.07-3.68; p < .05) of support more likely to be readmitted.^25^ However, our study differed as they examined social support through additional factors such as living situation, family contact, and time spent alone at home. Additionally, the follow-up period used in their study was the time to first HF readmission, not 30 days. Finally, Rørth et al.’s study in 2018 found that not being employed was associated with a higher risk of HF readmission, which contradicts our findings of no significance. Their study, with a follow-up period of 5 years, revealed that not being part of the workforce had a HR of 1.09 (95% CI 1.05-1.14) in relation to HF readmission risk.^26^

The final aim of this paper was to use a qualitative approach to explore the healthcare-associated events that occurred between a patient’s index HF admission and subsequent readmissions within a one-year time frame. In addition, this exploration exercise was to ascertain if linked population-based data can inform a deeper understanding of the specific events (or lack thereof) that contribute to readmissions. While hospital care is appropriate for severe HF cases, well-managed and coordinated care in the community can help maintain HF patients in stable condition, which reduces the need for readmissions.^11^ According to the Australian Commission on Safety and Quality in Health Care, a multifaceted approach is necessary to decrease HF hospitalisations; this includes primary prevention, empowering patients, utilizing medications more effectively, increasing exercise and cardiac rehabilitation programs, and improving community care with better integration with hospital care and access to multi-disciplinary care.^12^

Figure 4 and Figure 5 above clearly illustrate a patient’s journey in the healthcare system. There was an increased number of interactions between health services for patients who had HF readmission compared to those who did not. However, no patterns or relationships could be drawn from this exercise. Additionally, the lack of clinical data further compounded the weakness of this approach. In essence, this qualitative time-series approach allowed us to examine the patient’s interactions with the healthcare system comprehensively. However, while highly descriptive, it did not necessarily provide meaningful insights into the complex interplay of events (or lack thereof) that contributed to readmissions. Further studies must be conducted to verify if a population-based dataset can be used qualitatively to develop targeted interventions to reduce the risk of readmissions and improve the care and outcomes for patients with HF.

### Strengths

In conjunction with the linked population-based dataset, a mixed-method research approach has allowed this paper to explore HF readmissions in a substantial breadth, i.e., quantitatively, and depth, i.e., qualitatively, which would not have been possible in a single-methods approach. One significant advantage of using linked population-based data is the ability to access a wide range of datasets. These datasets have enabled the prospective exploration of numerous determinants influencing HF readmissions, including patient health and sociodemographic characteristics. Moreover, the linked population data set is advantageous because it did not restrict the study to hospitalisation records but allowed us to investigate events occurring outside the hospital. Another advantage is the use of the 45 and Up Study dataset. A compelling feature of the 45 and Up Study is the large sample size of participants.^20^ Additionally, the APDC dataset for the 45 and Up study includes all records of hospital separations for all residents of NSW in both public and private hospitals, resulting in no selection biases when identifying our study participants.

### Limitations

The limitations of this study stem from the accuracy of the record linkage, the reliability of each dataset, the definitions used, and the validity of hospital records coding.

Even though the 45 and Up cohort embodies a broad representation of the Australian population, it was acknowledged that there was oversampling, specifically those aged 80 years and above who reside in rural or remote areas. Additionally, the cohort were potentially healthier and may have had lesser hospitalisation than the general population of the same age group.^20^ Moreover, the baseline recruitment had a participation rate of about 18.0%; thus, the cohort may differ slightly from the general population. Additionally, exposure data utilised in this research were primarily obtained from self-reports through the 45 and Up Study.

Additionally, exposure data were not synchronised with the time of hospital admission or readmission; the time elapsed between survey data and the index admission varied among patients. Another drawback was the lack of clinically significant factors unavailable through administrative data, which may affect HF readmission rates. These include but are not limited to the severity of HF, clinical parameters, diagnostic and pathology results, management plans and compliance with medications or follow-up procedures. HF research is challenging since it does not encompass a single, clearly defined disease but rather a complex collection of symptoms. As a result, this paper acknowledges that there may be ambiguity regarding the diagnosis and reporting of HF in hospital separation data.

## Conclusion

HF and its associated readmissions have been an ongoing public health issue with a substantial burden on the health system, and improvements to reduce this are long overdue. Our study found that the older-old adults, non-compliance with alcohol intake guidelines, and having one or more chronic conditions are significant risk factors for readmission. These findings suggest that there is a need for interventions to reduce alcohol intake and improve the management of comorbidities in older adults with HF. The quantitative findings of this study can be used to develop strategies for preventing HF readmission. Nevertheless, further population-linked studies, with clearly defined HF diagnosis and standardisation of readmission definition, are needed for a more accurate description of the burden of HF and its associated readmission in Australia. The qualitative findings did not provide meaningful insights into the complex interplay of events (or lack thereof) that contributed to readmissions. Further studies must be conducted to verify if a population-based dataset can be used qualitatively to develop targeted interventions to reduce the risk of readmissions and improve the care and outcomes for patients with HF.

## Data Availability

Data may be obtained from a third party and are not publicly available. Data that support the findings of this study are available from the Sax Institute, but restrictions apply to the availability of these data, which were used under license for the current study and so are not publicly available. The data, however, are available from the authors upon reasonable request and with permission from the Sax Institute

## Acknowledgement

The research was conducted using data retrieved from the 45 and Up Study (www.saxinstitute.org.au/our-work/45-up-study/). The 45 and Up Study is managed by the Sax Institute in partnership with partners Cancer Council NSW and NSW Ministry of Health. The authors express their gratitude to the many thousands of participants in the 45 and Up Study. The authors acknowledge the contributions of the NSW CHeReL for their role in data linkage (http://www.cherel.org.au/) and Service Australia for the provision of the MBS data and PBS data. The authors also acknowledge the SURE for providing a secure data access.

## Abbreviations

ABS: Australian Bureau of Statistics
aHR: Adjusted Hazard Ratio
AIHW: Australian Institute of Health and Welfare
APDC: Admitted Patient Data Collection
BMI: Body Mass Index
CHeReL: Centre for Health Record Linkage
CIs: Confidence intervals
DSSI: Duke Social Support Index
HF: Heart Failure
HR: Hazard Ratio
ICD-10-AM: International Statistical Classification of Diseases and Related Health Problems, 10th Revision, Australian Modification
MBS: Medicare Benefits Schedule
NSW: New South Wales
PBS: Pharmaceutical Benefits Scheme
RBDM: NSW Registry of Births, Deaths and Marriages
SEIFA: Socio-Economic Indexes for Areas

